# EFFECTIVENESS OF ACAMPROSATE FOR TINNITUS: A SYSTEMATIC REVIEW AND META-ANALYSIS OF CLINICAL TRIALS

**DOI:** 10.1101/2021.06.09.21258585

**Authors:** Osmar Clayton Person, Fernando Veiga Angélico Júnior, Rodrigo Lima de Godoy Santos, William Marasini de Rezende, Maria Fernanda Giusti, Maria Eduarda dos Santos Puga

## Abstract

**Introduction:** The effectiveness of tinnitus treatment represents a huge gap in the medical science. Acamprosate is a glutamatergic antagonist drug and GABA-agonist that could be used to control tinnitus due to its action on peripheral and central neurotransmission.

**Purpose:** To assess the effectiveness of acamprosate in the treatment of tinnitus.

**Material and Methods:** This is a systematic review and we searched for randomized clinical trials linking acamprosate to tinnitus in six databases: Cochrane - Central Register of Controlled Trials - CENTRAL (2021), PUBMED (1966-2021), EMBASE (1974-2021), IBECS (1982-2021), QINSIGHT (2021) and SCOPUS (2021). Two researchers independently extracted the data and assessed the quality of the studies.

**Results:** Two trials involving 121 patients were included. The methodological quality of these studies was low. Both studies evaluated as primary outcome the efficacy of acamprosate in improving tinnitus. The meta-analysis by random model resulted in no significant difference between the groups treated with acamprosate and placebo (RR = 3,69, 95% CI 0,87-15,62; p=0,08), considering tinnitus improvement. **Conclusions:** There is no evidence that acamprosate is effective for tinnitus treatment. We recommend new trials using rigorous methodology. Randomization and blinding should be of the highest quality, given the subjective nature of tinnitus and the strong likelihood of a placebo response. The CONSORT statement should be used in the design and reporting of future studies.

## BACKGROUND

Tinnitus corresponds to the perception of sound in the absence of external acoustic stimulation, which may have low impact or even represent a debilitating condition to the wearer.^1^ Up to 18% of the population of industrialised societies are affected by tinnitus and 0.5% indicate that the symptom severely affects their quotidian.^2^

For more than 50 years, Heller and Bergman have demonstrated that any normal person placed in a sufficiently silent environment can feel sounds inside their head, assuming that tinnitus activity is a phenomenon naturally perceived by many in environments under these conditions.^3^

However, currently, no specific therapy has been recognized as satisfactory in all patients who suffer from tinnitus, and not even widely disseminated therapeutic options in the media and society, as for example the use of Ginkgo biloba4 extract and Zinc^5^ showed efficacy in secondary studies with the best level of evidence available in the world literature.

Azevedo & Figueiredo (2005) were pioneers in describing that the oral administration of acamprosate could be effective in the therapeutic approach of tinnitus. Since then, the world scientific community has slowly considered this possibility.^6^

Tinnitus very possibly has its generator mechanism related to neurotransmitters and neurotransmission. In the auditory pathway afferent the presence of glutamate and gamma-aminobutyric acid (GABA) is well described, while in the efferent pathway, in addition to these, dopamine and acetylcholine are known.^7^

Glutamate is the most prevalent neurotransmitter in the nervous system and it exerts a excitatory activity, while GABA is synthesized from the glutamate itself through the enzyme glutamate-decarboxylase, acting inhibitory in the synapses.^8^

The Acamprosate is a synthetic drug used in the treatment of alcoholism. Its mechanism of action involves both glutamatergic and GABA-ergic system.^9^ This drug reduces the action of glutamate in the central nervous system, particularly its excitatory action in NMDA (N-methyl-D-aspartate) receptors, possibly through the blockade of calcium channels. It also increases the number of recapture sites of GABA, broadening the GABA-ergic neurotransmission and thus inhibiting the excitatory activity in the auditory pathways.^10,11^ No other drug routinely used in the therapeutic approach of Tinnitus acts concomitantly in excitatory and inhibitory systems, which allows considering that, in theory, acamprosate could aid the treatment of tinnitus.6

The possibility of this drug improving tinnitus is considered in the medical community^9^, especially among otorhinolaryngologists. Much is commented in the media that acamprosate constitutes a very useful medicine in the treatment of tinnitus, but the scientific support lacks evidence. Given this evidentiary scientific gap and the search for the best evidence available in the literature, we proposed the development of this study.

### OBJECTIVES

The purpose of this study was to evaluate the effectiveness of acamprosate in the treatment of subjective tinnitus in adults, especially in relation to:

- The subjective clinical improvement of the symptom;
- Improvement in quality of life;
- Adverse effects on pharmacotherapy.

## MATERIALS AND METHODS

This is a systematic review of randomized clinical trials, following the methodology recommended by the Cochrane collaboration^12^.

Only randomized (RCT) and quasi-randomized clinical trials were included in the study, whose participants were adults of both sexes with unilateral or bilateral subjective tinnitus, regardless of the severity and time of the symptom.

### Types of Interventions

Group treated with acamprosate, regardless of dosage or time of treatment, compared with placebo-treated group.

### Types of Outcomes

- ***Primary***
  - Subjective tinnitus improvement

- ***Secundary***
  - Improved quality of life
  - Adverse effects

### Search Methods for Identification of Studies

The search strategy developed was the one recommended by chapter 6 of the Cochrane Collaboration Handbook, a high sensitivity search strategy. The keywords “tinnitus” and “acamprosate” were used, also using the Cochrane filter to identify the studies (randomized clinical trial - RCT).

Six electronic databases were searched: Cochrane Library - CENTRAL (2021), MEDLINE/PUBMED (1966-2021), EMBASE (1974-2021), IBECS (1982-2021), QINSIGHT (2021) and SCOPUS (2021). The date of the last survey was April 7, 2021. There were no restrictions on the language or geographic origin of the publications.

## Data Collection and Analysis

The citations obtained through the search strategy in the various databases were gathered in a single list, after excluding the duplicate citations. The titles and abstracts of all studies were reviewed and those considered to be potentially relevant were selected for full reading. Those who fulfilled the selection criteria were included in the review. The entire selection process of the studies was performed in pairs by two independent reviewers.

Both independently extracted the relevant data from each selected study for inclusion and compared its findings. For each study, information was collected regarding the characteristics of the study, the participants, the interventions and the outcomes.

The methodological quality of the included studies was also evaluated by two independent researchers, according to the recommendations of the Cochrane Handbook.^12^

Each RCT received a final score for each of six domains, according to the overall risk of bias (Table 1), being considered: YES (low risk of bias), UNCERTAIN (risk of uncertain bias) or NO (high risk of bias), where:

**Table 1.** Analysis of bias risk.

| DOMAIN | DESCRIPTION | TRIAL |
| --- | --- | --- |
| **Generation of the randomization sequence** | Description of the method used to generate the allocation sequence in sufficient detail to enable the evaluation to result in comparable groups | Was the allocation sequence properly generated? |
| **Allocation concealment** | Description of the method used for the allocation concealment of the sequence with sufficient details to determine whether the allocation of the intervention could be known between the time of randomization and the administration of the intervention | Was the allocation concealment adequate? |
| **Blinding of participants and researchers** | Description of all the measures used to maintain the blinding of participants and researchers until the end of the study. Provides some information if the blinding was effective. | Was the knowledge of the intervention allocation adequately prevented during the study? |
| **Incomplete data on outcomes** | Description of all outcome data, including losses and exclusions in the analysis. When there are losses and exclusions, it describes the number in each intervention group and reasons. | Were the follow-up losses adequately reported and analyzed? |
| **Selective reporting of outcomes** | There is a possibility of selective reporting of some of the pre-specified outcomes. | Are the results of the study free from a selective report of outcomes? |
| **Other sources of bias** | Description of any questions about possible biases not previously analysed | Is the study seemingly free of other problems that can lead to the risk of bias? |

- Low risk of systematic error or bias: All criteria well described and appropriately applied;
- Uncertain risk of systematic error or bias: one or more of the first three criteria could not be evaluated due to lack of information for the trial;
- High risk of systematic error or bias: one or more of the first three criteria inappropriately applied.

## Statistical analysis

The data analysis was performed comparing the outcomes of interest between the acamprosate and placebo treated groups. Comparable data were analyzed using the software *Review Manager 5*.*3*.^13^

As the outcomes in analysis involved dichotomous variables, the risk difference (RD), relative risk (RR) and respective confidence intervals of 95% (95%CI) were calculated. The relative risk is the risk ratio between the group treated with acamprosate and the control group (placebo or other treatment); an RR greater than 1 is indicative of favorable outcome (improvement in tinnitus).

The RD is the absolute risk reduction of the acamprosate-treated group over the placebo-treated control.

The unit of analysis was the individual patient.

### Heterogeneity assessment

The heterogeneity was evaluated through the chi-square test with N degrees of freedom, N being equal to the number of studies that contributed with the data, minus one. To quantify the inconsistencies between the sum estimates, the I^2^ test (I^2^ = [(Q – gl)/Q] x 100%), where “Q” is the chi-square statistic and “gl” its degrees of freedom, was used. Values of I^2^ greater than 50% were considered indicative of substantial heterogeneity.^121^

It was foreseen, in the hypothesis of no significant heterogeneity, that combined estimates of the treatment effect were computed for each result, using a fixed-effect model. In the condition of significant heterogeneity, the use of the random effect model was predicted.

## RESULTS

The search strategy recovered April 2021 a total of 59 citations, 3 in Cochrane, 9 in PUBMED, 6 in EMBASE, 2 in IBECS, 27 in QINSIGHT and 12 in SCOPUS. After elimination of duplicate citations (n = 15), there were 44 unique studies. After reading the titles and summaries of these studies, 37 were excluded because they did not meet the selection criteria and 7 were selected for reading, after which two met the criteria and were included in this systematic review (Figure 1).

**Figure:**
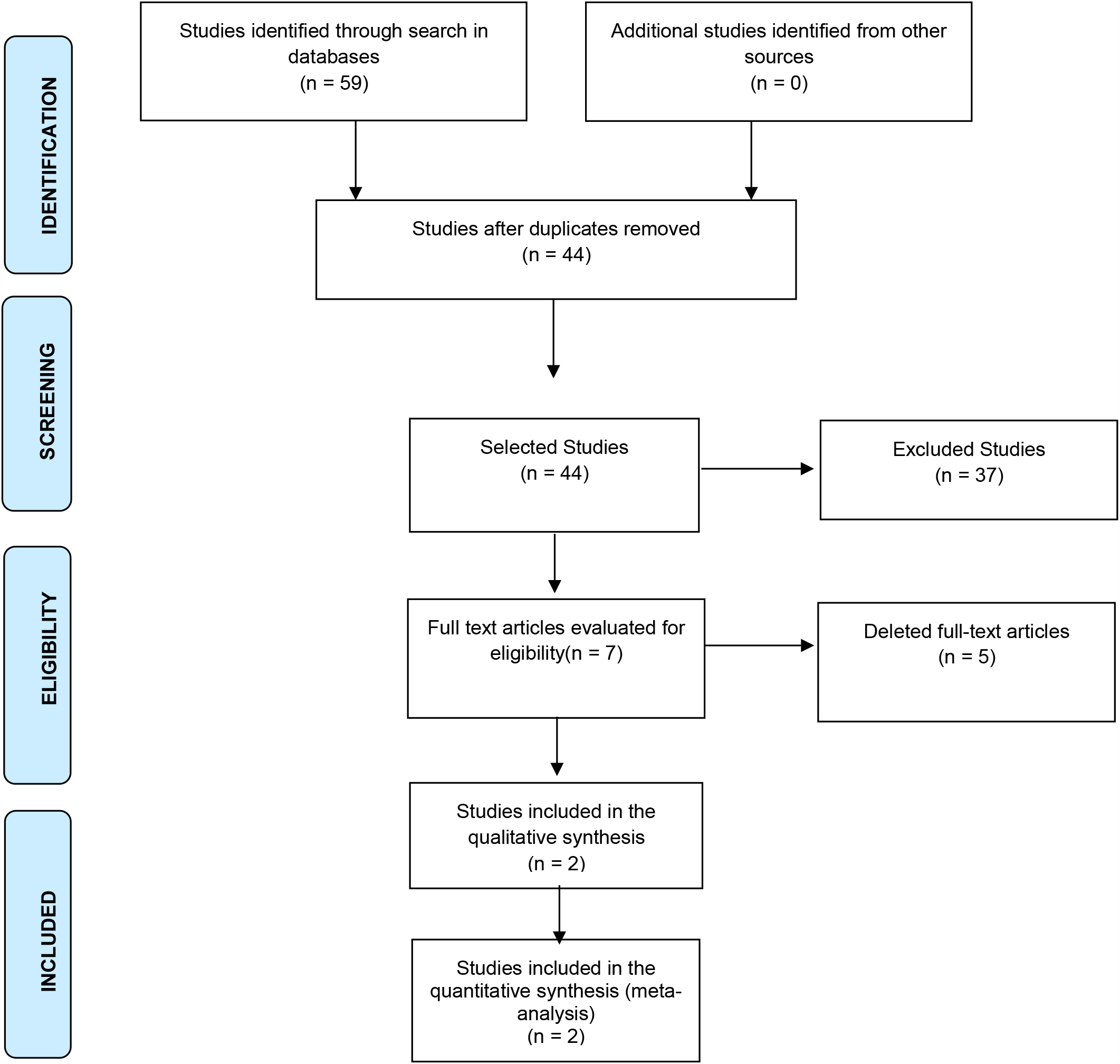
Flowchart of the process of identification of studies in electronic databases.

### Included Studies

Two studies^6,9^ with a total of 121 participants were included in the systematic review. The main characteristics of these studies are in Table 4. The studies were published in 2005 and 2012, respectively, in Brazil and India.

### Characteristics of the Included Studies

The study of Azevedo and Figueiredo (2005)^6^ was conducted in a double-blind randomized clinical trial (RCT), conducted in Volta Redonda, Rio de Janeiro, Brazil, and involved 50 patients with sensorineural tinnitus who constituted two groups of 25 patients. Patients with alterations in the external ear and middle ear, as well as those with conductive or mixed dysacusis were excluded from the study.

The first group received treatment with acamprosate (333 mg, three times a day) for 90 days, and the second group was treated with placebo for the same period.

Before treatment began and after 30, 60 and 90 days of treatment, the patients were evaluated by an analogue scale, assigning a score of zero to ten (with half at half point fractionation) to their tinnitus, related to the degree of discomfort.

Statistical analysis was performed using Student’s t-test for independent samples or the Mann-Whitney test. To evaluate the evolution of the tinnitus scale, we performed the Friedman Variance Analysis.

There was a loss of 18% of the patients, being 2 (4%) in the Acamprosate Group and 7 (14%) In the placebo group. 6 patients (1 from the Acamprosate Group and 5 from the placebo group) discontinued the medication due to adverse effects and 3 patients due to family pressures.

The authors described a significant reduction in the score attributed to the numerical scale of tinnitus discomfort in the group treated with acamprosate over time of medication use (p ‹ 0.0001). There was no difference in the placebo-treated group (P = 0.22).

The authors did not evaluate any improvement in the quality of life of the patients.

Mild adverse effects were reported, with no difference between the groups treated with acamprosate and placebo (p = 0.35). In the group treated with acamprosate the adverse effects were mild (choking and epigastralgia) and in one case the patient developed depression, whose relation with the use of the drug was not clear.

The study by Sharma (2012)^9^ was in a double-blind randomized clinical trial (RCT) crossover, conducted in Punjab (India) and involved 45 patients with sensorineural tinnitus. Patients who were pregnant or breastfeeding, those with alterations in the external ear and middle ear, as well as those with conductive or mixed dysacusis or a history of barotrauma were excluded from the study.

The patients were divided into two groups, the first was treated with acamprosate (333 mg, three times a day) for 45 days, and the second group was treated with placebo for the same period. After 45 days, a seven-day washout period was given and the groups received the opposite treatment.

The patients were evaluated by an analog scale, assigning a score from zero to ten to assess the degree of tinnitus discomfort.

Regarding tinnitus improvement, there was a reduction in the symptom in 92.5% of patients treated with acamprosate, while only 12.5% of those treated with placebo. The author has not provided statistical details.

An evaluation of the improvement in patients’ quality of life was carried out through a validated questionnaire, which showed an improvement in the quality of life of patients treated with acamprosate compared to placebo. The mean scores on the questionnaire were 42.33 in the acamprosate group and 67.19 in the placebo group. Statistical analysis was not reported.

The author described no adverse effects in the study in any of the groups. There were 11.1% of patients lost, 2 (4.4%) felt worse and received another treatment and 3 (6.7%) gave up the study. The author did not provide further details.

Table 2 presents the characteristics of included studies.

**Table 2.** Characteristics of included studies.

| Study | Method | Participants<br>(both gender) | Treatment | Duration | Primary<br>Outcomes |
| --- | --- | --- | --- | --- | --- |
| Azevedo<br>2005<sup>6</sup> | RCT | Included: 50<br>Analyzed: 41 | 333 mg<br>acamprosate<br>(3x/day) x<br>placebo<br>(3x/day) | 90 days | Tinnitus<br>improvement<br>Safety of the<br>medicinal<br>product |
| Sharma<br>2012<sup>9</sup> | RCT<br>(Crossover) | Included: 45<br>Analyzed: 40 | 333 mg<br>acamprosate<br>(3x/day) x<br>placebo<br>(3x/day) | 45 days | Tinnitus<br>improvement<br>Changes in<br>quality of life<br>Safety of the<br>medicinal<br>product |

### Risk of bias

Table 3 presents the bias risk analysis of the three RCTs included in this systematic review.

**Table 3.** Analysis of bias risk in the RCTs included.

| ***TRIAL (RISK)*** | ***AZEVEDO***<br>***(2005)*** | ***SHARMA (2012)*** |
| --- | --- | --- |
| ***Randomization***<br>***sequence*** | UNCLEAR | UNCLEAR |
| ***Allocation concealment*** | UNCLEAR | UNCLEAR |
| ***Blinding of participants and***<br>***researchers*** | UNCLEAR | UNCLEAR |
| ***Description of***<br>***losses in follow-***<br>***up and analysis*** | YES | YES |
| ***Selective reporting of***<br>***outcomes*** | YES | YES |
| ***Other sources of bias*** | YES | YES |

### Meta-analysis

The available data allowed the statistical processing for the primary outcome (improvement of tinnitus), which is presented in a forest graph (Figure 2).

**Figure 2:**
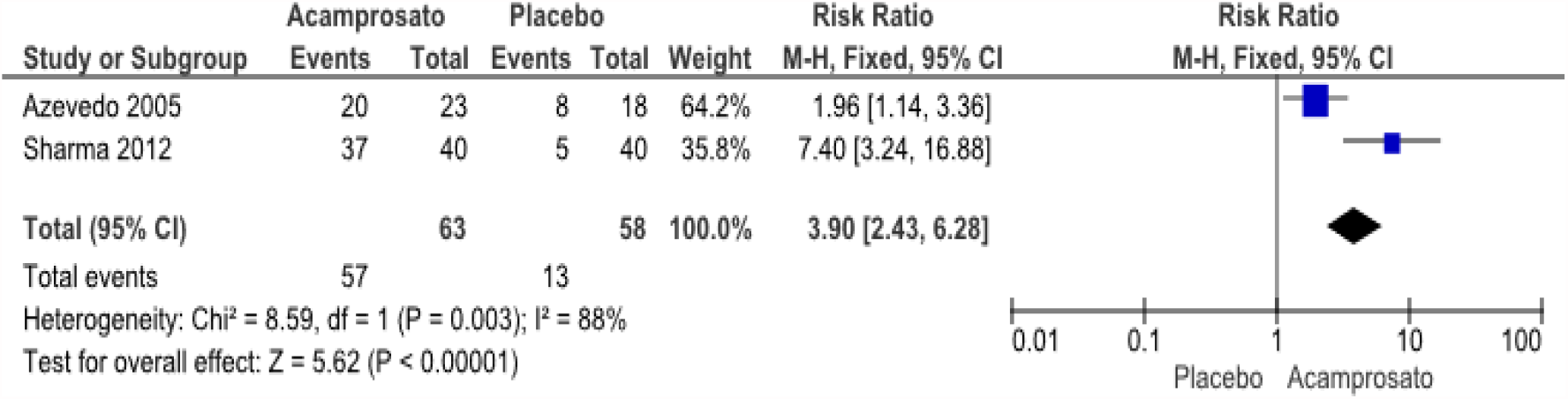
Meta-analysis related to tinnitus improvement with acamprosate versus placebo considering fixed effect model.

The statistic was favorable to treatment with acamprosate (RR = 3.90 - CI95% 2.43-6.28; p <0.00001). However, the heterogeneity showed to be quite high (I^2^ = 88%).

Due to the high heterogeneity (I^2^ = 88%) found in the fixed effect model, we opted for statistical processing in a random effect model - Figure 3.

**Figure 3:**
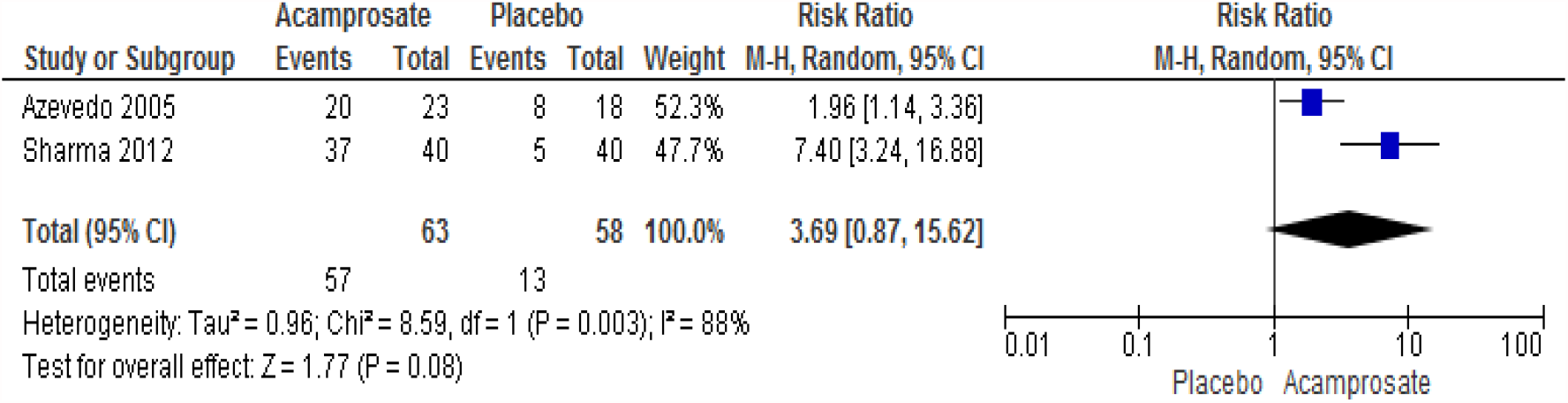
Meta-analysis related to tinnitus improvement with acamprosate versus placebo considering random effect model.

## DISCUSSION

Intra and extracellular ionic concentration differences are fundamental in the process of depolarization and synaptic transmission. Although many ions participate in this process, the chlorine and calcium ions play a very relevant role. The post-synaptic receptors of GABA-ergic fibers allow the chlorine to enter inside the neuron, inducing hyperpolarization and reducing the conduction of electrical potential. In the opposite line, the glutamatergic post-synaptic ionotropic receptors, especially those of the NMDA type, allow the entry of calcium into the cell, inducing depolarization and facilitating the transmission of the potential. Several studies have sustained the association of glutamate-GABA imbalance to the modulation of sensorineural tinnitus.^14^

Moreover, the excess of glutamate released in the synaptic cleft generates excitotoxicity, which can injuriate the neuron by the excess of calcium in the intracellular medium. Excitotoxicity induces overactivation of NMDA postsynaptic ionotropic receptors, and this is a widely accepted pathophysiological mechanism among researchers as a tinnitus generator.^14,15^

Acamprosate, due to its mechanism of peripheral and central action in Glutamatergic and GABA-ergic systems, especially with regard to inhibition of glutamic uptake in postsynaptic NMDA receptors, in a thesis, could contribute to the reduction of tinnitus sensation.^5,14,15^

This systematic review of the literature identified only two RCTs, which evaluated the efficacy of acamprosate for tinnitus assuming as control a group of patients treated with placebo. A strong point of this review was its methodological rigor, following the recommendations of the Cochrane collaboration, including a sensitizing and unrestrained search, the involvement of two independent investigators in the selection of studies, extraction of and evaluation of the quality of the included studies. One limitation was the fact that it did not search for unpublished studies on the subject, possibly described in abstracts of congresses, symposiums and scientific journeys. Also, no search was conducted with the pharmaceutical industry. Currently, tinnitus is considered an incurable symptom.^5^ The clinical manifestations associated with it are quite variable, with patients with a similar tinnitus pattern observed at acuphenometry, but with a totally distinct behavior regarding the characterization of the nuisance.^6^

In the 1990s, Jastreboff^16,17^ described tinnitus as a symptom originating mainly in the peripheral auditory system, especially in the cochlea. However, he considered that the acoustic signal could suffer interference from extra-auditory systems, especially the limbic system, as it ascends to the subcortical areas.

Thus, in an individual with tinnitus, it could occur habituation to the symptom, whose clinical repercussion regarding the discomfort would be very. low or even null. In other people, the limbic recruitment, associated with anxiety, tension, fear and correlation with negative feelings, would result in a summation effect, with consequent perception of intense and persistent tinnitus. This neurophysiological model contributes singularly to the comprehension of the relevance of tinnitus in the daily life of the individual, which warns us about the subjectivity and difficulty of characterizing eventual clinical responses to the proposed drug treatments.^17^

As for the evaluation of patients with tinnitus, the use of self-assessment questionnaires of the symptom, such as the THI (Tinnitus Handicap Inventory), is presented as a good predictor of patients with a high degree of anxiety and depression^18^, assisting in the diagnostic amplitude and consequent global therapeutic approach, which should always be individualized.^19^

Tinnitus treatment involves the proper diagnosis and the standardization of questionnaires can effectively contribute to the adequacy of each situation to the therapeutic reality.^19^ It is important to highlight that in the last two decades, the therapeutic approach of tinnitus has undergone modifications in proportion close to the new discoveries in the area. In this century, the new pathways and. connections related to the potentialization of the symptom, such as the binding of trigeminal innervation in the face^20,21^, should trigger new therapeutic processes.

The importance of the somatosensory system in the perception of tinnitus regarding the modulation of the symptom by means of rapid maneuvers, such as cervical movement or bite, has been emphatically described in the literature.^20,21^ The patients report reduction or increase of the tinnitus during the maneuvers, linking somatic modulation to tinnitus at the level of the referred sensation.

It is known, in the literature, that about 300 different diseases and conditions may be associated with tinnitus.^22^ In this context, the correlation with underlying disease plays a prominent role in the treatment, as the reversal of the disease should be a priority, whenever possible, even aligned with the possibility of easing the sound sensation.

Therefore, it is lawful to consider that tinnitus is exposed in multiple scenarios, and the allocation of symptomatic patients in subgroups allows greater homogeneity, such as patients without hearing loss and patients with cochleopathy and complaint of Hypoacusis or presence/absence of signs of limbic recruitment.

Historically, the world literature does not present many studies with a good level of evidence that allow conclusions with solidity. Most therapies that. aim to improve tinnitus and consequently the quality of life are based on case reports and expert experience. These are empirical conducts, without clinical-scientific support, or permeated in proven concepts only in in vitro experiments or in experimental animals, but until then not demonstrated regarding the effectiveness in humans.^23^

Although the constructivism of science is progressive, in recent decades most RCTs for the treatment of tinnitus have been financed by the pharmaceutical industry which, in the light of the market, outlined the marketing interests of one or the other medicinal products. It was the case of the extract of Ginkgo Biloba, launched in the market at high prices for the consumer, as being effective for the treatment of many potentially pathological conditions, among which tinnitus.

Even among the current tinnitus-related RCTs, few have been conducted considering the diversity of cases under the foundation of the same symptom.

Subject of this systematic review, acamprosate was evaluated in only two studies with placebo control. The number of participants in these studies was very small and the heterogeneity found in the very high meta-analysis (I^2^= 88%), which evidences limitation in the interpretation of real effectiveness in the treatment of tinnitus.

These are low methodological quality RCTs and statistical processing suggests that the treatment with acamprosate is more effective than placebo in the clinical improvement of tinnitus (figure 2), but maximum caution is required. Statistical analysis using a fixed-effect model shows that acamprosate favors placebo (RR = 3.90 - 95% CI 2.43-6.28). However, in due to high heterogeneity (I^2^ = 88%), it was necessary to perform statistics using a random effect model (Figure 3) and we found that there is no difference between groups (RR=3,69 IC95% 0,87-15,62; p=0,08). The evidence is very low and very limited. Only mild adverse events are described, most commonly related to gastric intolerance.

In the context, it is essential to perform new RCTs that involve the therapy with acamprosate for patients with tinnitus, and other parameters should be considered by the researchers in light of the recent discoveries of science. The new studies should involve the allocation of patients with tinnitus in subgroups, as much as possible, according to cause, pattern of hearing loss according to audiometry and existence of signs of recruitment. The use of standardized and validated questionnaires, such as THI is very important, as well as the rigorous methodological standard, which should cover the detailed description of randomization and blinting of patients and investigators. It is also highly recommended to report in accordance with Cochrane’s CONSORT Statement.

The multiple events associated with tinnitus and highlighted here, including signs of anxiety and depression, inferring the limbic system, different patterns of dysacusis and associations with metabolic disorders and musculature of the face, especially in the area of sensory innervation of the trigeminal nerve, minimally suggest the need of inclusion of the patients in subgroups of studies, considering the use of standardized and validated questionnaires to evaluate the outcomes of interest. In addition, the specificities related to the test treatment should be considered in future studies.

## CONCLUSION

There are few studies on the effectiveness of acamprosate for treating tinnitus. The sampling in these studies is small and there is substantial heterogeneity in these studies. The level of evidence is very low and these findings recruit the urgent need for new randomized clinical trials with rigorous methodological quality, detailed description of the randomization and blinding method of the participants, and the use of standardized and validated tinnitus questionnaires. It is also very important to report the new RCTs in accordance with the CONSORT Statement.

## Data Availability

does not apply because it is a Systematic Review

## Notes

### Competing Interest Statement

The authors have declared no competing interest.

### Funding Statement

The Authors there was no financing

### Author Declarations

Universidade Santo Amaro UNISA Sao Paulo Brazil

